# Immunogenicity of COVID-19 Vaccination in Immunocompromised Patients: An Observational, Prospective Cohort Study Interim Analysis

**DOI:** 10.1101/2021.06.28.21259576

**Authors:** Ghady Haidar, Mounzer Agha, Amy Lukanski, Kelsey Linstrum, Rachel Troyan, Andrew Bilderback, Scott Rothenberger, Deborah K. McMahon, Melissa Crandall, P. Nathan Enick, Michelle Sobolewksi, Kevin Collins, Marc B. Schwartz, Jeffrey M. Dueker, Fernanda P. Silveira, Mary E. Keebler, Abhinav Humar, James D. Luketich, Matthew R. Morrell, Joseph M. Pilewski, John F. McDyer, Bhanu Pappu, Robert L. Ferris, Stanley M. Marks, Cynthia Klamar-Blain, Urvi M. Parikh, Amy Heaps, Paula L. Kip, Alan Wells, Tami Minnier, Derek Angus, John W. Mellors

## Abstract

**Objectives:** Immunocompromised patients were excluded from COVID-19 vaccine clinical trials. The objectives of the study were to measure antibody responses, levels, and neutralization capability after COVID-19 vaccination among immunocompromised patients and compare these variables to those of immunocompetent healthcare workers.

**Methods:** This is an interim analysis of an ongoing observational, prospective cohort study which launched on April 14, 2021 across Western Pennsylvania. Participants were healthy healthcare workers (HCW) and immunocompromised patients who had completed their COVID-19 vaccination series. Individuals with a history of COVID-19 were not eligible. Serum was collected to measure for the presence of IgG against the SARS-CoV-2 Spike protein using a semi-quantitative assay; antibody levels were available for comparisons. A quasi-random subset of patients was selected for pseudovirus neutralization assays. Seropositivity with 95% Clopper-Pearson exact confidence intervals and distribution of antibody levels were measured. To identify risk factors for seronegativity, clinical characteristics were univariately compared between antibody reactive and non-reactive individuals within the immunocompromised group.

**Results:** 107 HCW and 489 immunocompromised patients were enrolled. Compared to HCWs, seropositivity was significantly lower (p<.001) among immunocompromised patients with Solid organ transplant (SOT), autoimmune, hematological malignancies, and solid tumors (HCW=98.1%; SOT=37.2%; autoimmune=83.8%; hematological malignancies=54.7%; and solid tumor=82.4%, p < 0.05). Over 94% of patients with Human Immunodeficiency Virus were seropositive. Among seropositive patients, antibody levels were much lower among SOT (4.5 [2.1,13.1], p=.020). Neutralization titers tightly correlated with antibody levels (Spearman r = 0.91, p < 0.0001).

**Conclusion:** Our findings demonstrate the heterogeneity of the humoral immune response to COVID-19 vaccines based on underlying immunosuppressive condition and highlight an urgent need to optimize and individualize COVID-19 prevention in these patients. These findings also have implications on public health guidance, particularly given revised Centers for Disease Control and Prevention recommendations permitting vaccinated individuals to abandon masking and social distancing in most settings. Future studies are warranted to determine assessment of cellular immunity, longitudinal measurement of immune responses, and the safety and efficacy of revaccination.

## INTRODUCTION

Immunocompromised patients are at risk for severe and protracted severe acute respiratory syndrome coronavirus 2 (SARS-CoV-2) infection with the potential to generate and spread multiply-mutated variants.^1,2^ These patients should therefore be prioritized for COVID-19 vaccination but were excluded from clinical trials evaluating the immunogenicity and efficacy of COVID-19 vaccines due to confounding comorbidities.^3–5^ Not surprisingly, recent studies in transplant, oncology, and rheumatology patients have demonstrated that COVID-19 vaccines elicit demonstrable antibody responses well below the 100% response rates seen in healthy volunteers in the reported phase 1/2 trials.^6–9^

Despite these emerging data, several unknowns persist, including the degree of the antibody response in seropositive immunocompromised patients, and whether antibodies from immunocompromised patients are capable of neutralizing SARS-CoV-2. To address these knowledge gaps, we implemented the COVID-19 Vaccination in the Immunocompromised Study (CoVICS). The objectives of the study were to measure antibody responses, levels, and neutralization capability after COVID-19 vaccination among immunocompromised patients and compare these variables to those of immunocompetent healthcare workers (HCW). We hypothesized that compared to HCW, seropositivity, antibody levels, and neutralization titers will be lower among immunocompromised patients based on underlying disease and iatrogenic immunosuppression. We present here an interim analysis of our findings.

## METHODS

CoVICS is an ongoing observational, prospective cohort study embedded in the electronic medical record (EMR) of adult patients who had completed their COVID-19 vaccine series. The study was approved by the University of Pittsburgh Institutional Review Board (Study ID: 21030056/HCC 21-032). Enrollment began on April 14^th^, 2021. To obtain serum from patients across the entire University of Pittsburgh Medical Center (UPMC) Health System, a customized SARS-CoV-2 IgG order set was built in the EMR that was only used by the study team and was not billed to the patient. Serum could be drawn at any of 16 UPMC hospital-based labs across Western Pennsylvania; and test results were shared with the patients. The study was advertised through social media, e-mail, and the UPMC EMR. Participants were enrolled online using a contact-free system for electronic consent.

### Participants

Eligibility criteria for the immunocompromised arm of CoVICS included any of the following medical conditions: solid organ transplant (SOT), hematological malignancies, solid tumors being treated with systemic or radiation therapy over the past 12 months, autoimmune or chronic inflammatory conditions being treated over the past 12 months, and Human Immunodeficiency Virus (HIV) infection. For the control arm, we enrolled healthy HCWs at UPMC. Participants with a known history of prior COVID-19 infection were excluded. All participants were required to have completed a two-dose mRNA vaccination (Moderna mRNA-1273 or Pfizer BNT162b2) or a single dose of Ad26.COV2.S (Johnson & Johnson) at least 14 days prior to testing.

### Data Collection and Outcomes

Study data were collected by the project coordinators and managed using the Research Electronic Data Capture (REDCap) hosted at the University of Pittsburgh.^10^ REDCap is a secure, web-based software platform designed to support data capture for research studies.^11^ Serum was collected from participants after completion of their COVID-19 vaccine series and processed at UPMC’s CLIA-88 accredited Central Lab. The primary outcome was seropositivity, defined as the proportion of patients who had a reactive SARS-CoV-2 Spike protein IgG result following vaccination. As secondary analyses, we compared antibody levels across patient groups, pseudovirus neutralization titers, and risk factors for seronegativity, defined as having a negative SARS-CoV-2 Spike protein IgG result following vaccination.

#### Antibody assays

Serum underwent antibody testing using the Beckman Coulter SARS-CoV-2 platform (Spike (S) receptor-binding domain (RBD) IgG).^12^ These results are expressed as extinction coefficient (signal/cutoff) ratios or “levels” and interpreted as positive (≥ 1.00), equivocal (> 0.80 to < 1.00), or non-reactive (≤ 0.80).^13^ The assay was run according to the manufacturer’s instructions.^14^ Test result interpretations were automatically shared with the study participants through UPMC’s EMR. For data analysis, reactive results were defined as positive (seropositive), and equivocal or non-reactive results were defined as negative (seronegative). Antibody levels were available for comparisons.

#### Pseudovirus neutralization assays

We evaluated SARS-CoV-2 pseudovirus neutralization titer levels in a subgroup of patients. SARS-CoV-2 pseudovirus (PSV) was generated in 293T cells by co-transfection of pFC37K-CMV-S, an enhanced expression plasmid encoding for codon-optimized full-length SARS-CoV-2 S (Wuhan-1 sequence containing D614G substitution) with the N-term HiBit tag removed, and pNL4-3.luc.R-E-mCherry-luciferase, an envelope deficient HIV-1 dual reporter construct that was cloned by recombination of the pNL.luc.R-E-plasmid (NIH AIDS Reagent Program) and the fully infectious pNL4-3 mCherry luciferase plasmid (Addgene).^15–18^ After harvest, PSV was centrifuged at 400xg for 10 minutes at room temperature (RT) and supernatant removed and filtered with a 0.45 micron syringe filter to remove producer cells. For neutralization assays, 104 293T-hACE2 cells were plated in 100 microliters (μL) media per well in 96 well white-wall white-bottom plates (Perkin Elmer) and incubated overnight at 37°C. Sera was diluted 1:3, then serially diluted 3-fold and incubated with 50μL of 1:10 PSV for 1 hour at 37°C. After incubation, media was removed from wells containing 293T-hACE2 cells and replaced with PSV/sera, and spinoculation was performed at 1,000xg for 1 hour at RT. Plates were then incubated for 48 hours at 37°C. After 48 hours, plates were analyzed for luciferase production by adding 100μL of BriteLite Plus reagent (Perkin Elmer), incubating at RT for 2 minutes, and reading on a Victor Nivo microplate luminometer (Perkin Elmer). Results are reported as the highest serum dilution that neutralizes >50% of the PSV termed the NT50.

### Statistical Methods

Descriptive statistics for the study sample are expressed as means, standard deviations, counts, and percentages. Baseline socio-demographic characteristics, seropositivity, and post-vaccine antibody levels were compared between immunocompromised and immunocompetent participants using two-sample student *t*-tests (or Wilcoxon rank sum tests in the case of skewed distributions) for continuous variables and chi-square tests for categorical variables. Within the immunocompromised group, these same characteristics were presented descriptively, stratified by immunocompromising condition.

Seropositivity along with 95% Clopper-Pearson exact confidence intervals were plotted for immunocompetent individuals and immunocompromised patients by condition (autoimmune disease, hematological malignancy, solid tumor, SOT, and HIV) and compared by chi-square tests. The distributions of antibody levels were plotted for healthy controls versus individual immunocompromised conditions. Compared to controls, the distributions of antibody levels among immunocompromised patients were compared by the Wilcoxon rank sum test.

As essentially all immunocompetent individuals were reactive (98%), socio-demographic and clinical characteristics were univariately compared between antibody reactive and non-reactive individuals within the immunocompromised group. Again, student *t*-tests or Wilcoxon tests were used for continuous variables with chi-square tests used for categorical variables.

For the subsets of individuals with pseudovirus neutralization quantification, antibody levels and corresponding neutralization titer levels were plotted for immunocompetent versus immunocompromised individuals. The Spearman correlation coefficient was estimated between antibody and neutralization titer levels given their potentially nonlinear relationship. Analyses were performed using Stata SE, version 16.1 (College Station, TX) assuming a significance level of α=0.05. Methods and results are reported in accordance with Strengthening the Reporting of Observational Studies in Epidemiology (STROBE) statement.^19^

## RESULTS

### Participants

During the interim analysis period (April 14, 2021 through June 25, 2021), 596 participants were enrolled, 107 were healthy HCW (18%) and 489 were immunocompromised patients (82%). The distribution of immunocompromising conditions was as follows (**Table 1**): 183 SOT (37.4%), 160 autoimmune conditions (32.7%), 75 hematological malignancy (15.3%), 37 HIV (7.6%), and 34 solid tumors (7.0%). Overall, the majority of participants (98.2%) received the mRNA-1273 (Moderna=48.5%) or BNT162b2 (Pfizer=49.7%) vaccine series. Compared to HCWs, immunocompromised patients were older (mean age for HCW=43.7, Patients=59.5, p<.001) and less likely to be female (HCW=72%, Patients=51.1%, p<.001). Median days from 2^nd^ vaccine dose to antibody level drawn was 78 (IQR 58-105) and was longer for HCW compared to immunocompromised patients (HCW median days=119, Patient median days=77, p <.001).

**Table 1.** Descriptive characteristics in healthcare workers and immunocompromised patients (N=596).

| Characteristic | Healthcare<br>Workers<br>(N=107)<br>(18.0%) | Immuno-<br>compromised<br>Patients<br>(N=489)<br>(82.0%) | Solid Organ<br>Transplant<br>(N=183)<br>(37.4%) | <u>Patients by Immunocompromised Condition</u> |  |  |  |
| --- | --- | --- | --- | --- | --- | --- | --- |
|  |  |  |  | Autoimmune<br>(N=160)<br>(32.7%) | Hematologic<br>Malignancy<br>(N=75)<br>(15.3%) | HIV*<br>(N=37)<br>(7.6%) | Solid<br>Tumor<br>(N=34)<br>(7.0%) |
| Age in years, mean (SD) | 43.7 (13.7) | 59.5 (13.7) | 61.2 (13.4) | 54.2 (14.8) | 66.3 (10.4) | 57.1 (10.1) | 63.1 (9.9) |
| Age in years, n (%) |  |  |  |  |  |  |  |
| 19 to 44 years | 62 (57.9%) | 79 (16.2%) | 24 (13.1%) | 47 (29.4%) | 2 (2.7%) | 3 (8.1%) | 3 (8.8%) |
| 45 to 60 years | 33 (30.8%) | 140 (28.6%) | 45 (24.6%) | 49 (30.6%) | 15 (20.0%) | 20 (54.1%) | 11 (32.4%) |
| >60 years | 12 (11.2%) | 270 (55.2%) | 114 (62.3%) | 64 (40.0%) | 58 (77.3%) | 14 (37.8%) | 20 (58.8%) |
| Sex, n (%) |  |  |  |  |  |  |  |
| Female | 77 (72.0%) | 250 (51.1%) | 73 (39.9%) | 112 (70.0%) | 38 (50.7%) | 3 (8.1%) | 24 (70.6%) |
| Male | 30 (28.0%) | 239 (48.9%) | 110 (60.1%) | 48 (30.0%) | 37 (49.3%) | 34 (91.9%) | 10 (29.4%) |
| Race, n (%) |  |  |  |  |  |  |  |
| Non-White, n (%) | 11 (10.3%) | 31 (6.3%) | 12 (6.6%) | 9 (5.6%) | 6 (8.0%) | 4 (10.8%) | 0 (0.0%) |
| White, n (%) | 96 (89.7%) | 458 (93.7%) | 171 (93.4%) | 151 (94.4%) | 69 (92.0%) | 33 (89.2%) | 34 (100%) |
| Days from second vaccine to<br>antibody sample, mean (SD) | 110.6 (43.0) | 74.1 (27.4) | 69.8 (25.9) | 79.3 (28.0) | 73.5 (26.2) | 75.3 (29.0) | 73.2 (30.7) |

\*HIV: Human Immunodeficiency Virus.

### Study Outcomes

#### Seropositivity

Compared to HCWs of whom 98.1% [93.4%-99.8%] were seropositive, seropositivity was significantly lower among persons with SOT (37.2% [30.1%-44.6%], p<.001), hematological malignancies (54.7% [42.7%-66.2%], p<.001), solid tumors (82.4% [65.5%-93.2%], p=.009), and autoimmune/chronic inflammatory conditions (83.8% [77.1%-89.1%], p<.001), (**Figure 1**). In contrast, there was no difference in seropositivity between healthy HCW and patients with HIV (94.6% [81.2%-99.3%], p = 0.26), all of whom were receiving antiviral therapy, had undetectable viral loads, and CD4 counts > 200 cell/mm^3^. There was no association between time from vaccination and seropsitivity.

**Figure 1.**
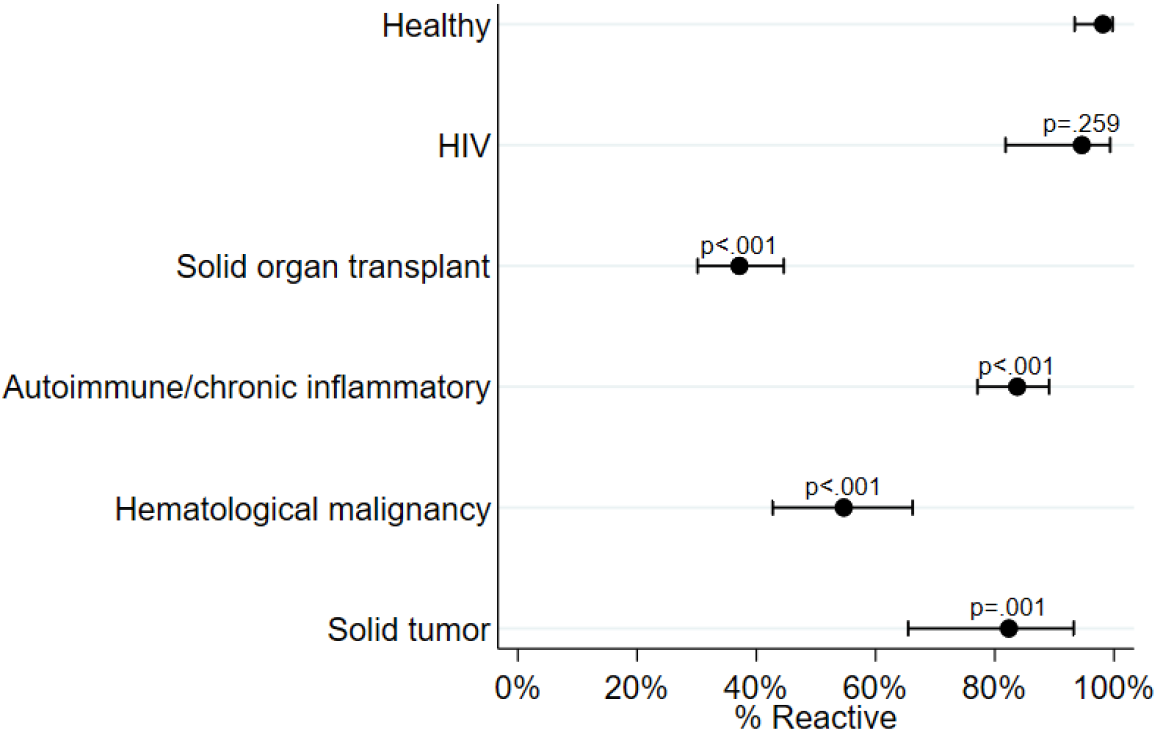
Seropositivity in healthy healthcare workers and immunocompromised patients. Comparisons between HCW and immunocompromised patients. Whiskers denote 95% confidence intervals. HIV, human immunodeficiency virus.

Type of vaccine was not associated with antibody response in either the HCW (both vaccines > 90%) or in the in the immunocompromised cohorts regardless of subgroup (seropositivity of 50.5% versus 49.5% for mRNA-1273 versus BNT162b2, respectively, p=0.427). Unadjusted risk factors for seronegativity after vaccination are shown in **Table 2**. Overall, age > 44 and male sex were associated with failure to produce SARS-CoV-2 antibodies. Among SOT recipients, lung transplant recipients had the lowest seropositivity (22.2%), but this was not statistically significantly different from the combined heart/liver/kidney group seropositivity of 38.8% (p=0.167). Liver transplant recipients in contrast had significantly higher seropositivity compared to other transplant types (60.6% vs 32.0%, respectively, p=0.002.) Additionally, use of an anti-metabolite and time from SOT (less than 1 year versus > 1 year) were associated with seronegativity. Similarly to the overall immunocompromised group, there was no statistically significant difference in the seropositivity by type of vaccine among SOT recipients (41.8% versus 33.0% for mRNA-1273 and BNT162b2, respectively, p=0.22).

**Table 2.** Seropositivity and antibody levels in healthcare workers and immunocompromised patients (N=596).

| **Outcome** | **Healthcare Workers**<br>**(N=107)** | **Immuno-compromised Patients**<br>**(N=489)** | **Solid Organ Transplant**<br>**(N=183)** | **<u>Patients by Immunocompromised Condition</u>** |  |  |  |
| --- | --- | --- | --- | --- | --- | --- | --- |
|  |  |  |  | **Auto-immune**<br>**(N=160)** | **Hematologic Malignancy**<br>**(N=75)** | **HIV\***<br>**(N=37)** | **Solid Tumor**<br>**(N=34)** |
| Seropositivity, n (%) | 105 (98.1%) | 306 (62.6%) | 68 (37.2%) | 134 (83.7%) | 41 (54.7%) | 35 (94.6%) | 28 (82.4%) |
| Beckman Coulter antibody level, mean (SD) | 10.1 (8.7) | 7.0 (9.6) | 3.4 (7.6) | 8.2 (8.3) | 8.6 (12.7) | 10.9 (8.6) | 12.6 (11.8) |
| Beckman Coulter antibody level, n (%) |  |  |  |  |  |  |  |
| 0 to < 5 | 40 (37.4%) | 296 (60.7%) | 151 (82.5%) | 78 (49.1%) | 46 (61.3%) | 10 (27.0%) | 11 (32.4%) |
| 5 to 10 | 26 (24.3%) | 70 (14.3%) | 13 (7.1%) | 31 (19.5%) | 7 (9.3%) | 13 (35.1%) | 6 (17.6%) |
| More than 10 | 41 (38.3%) | 122 (25.0%) | 19 (10.4%) | 50 (31.4%) | 22 (29.3%) | 14 (37.8%) | 17 (50.0%) |

\*HIV: Human Immunodeficiency Virus.

**Table 3.**
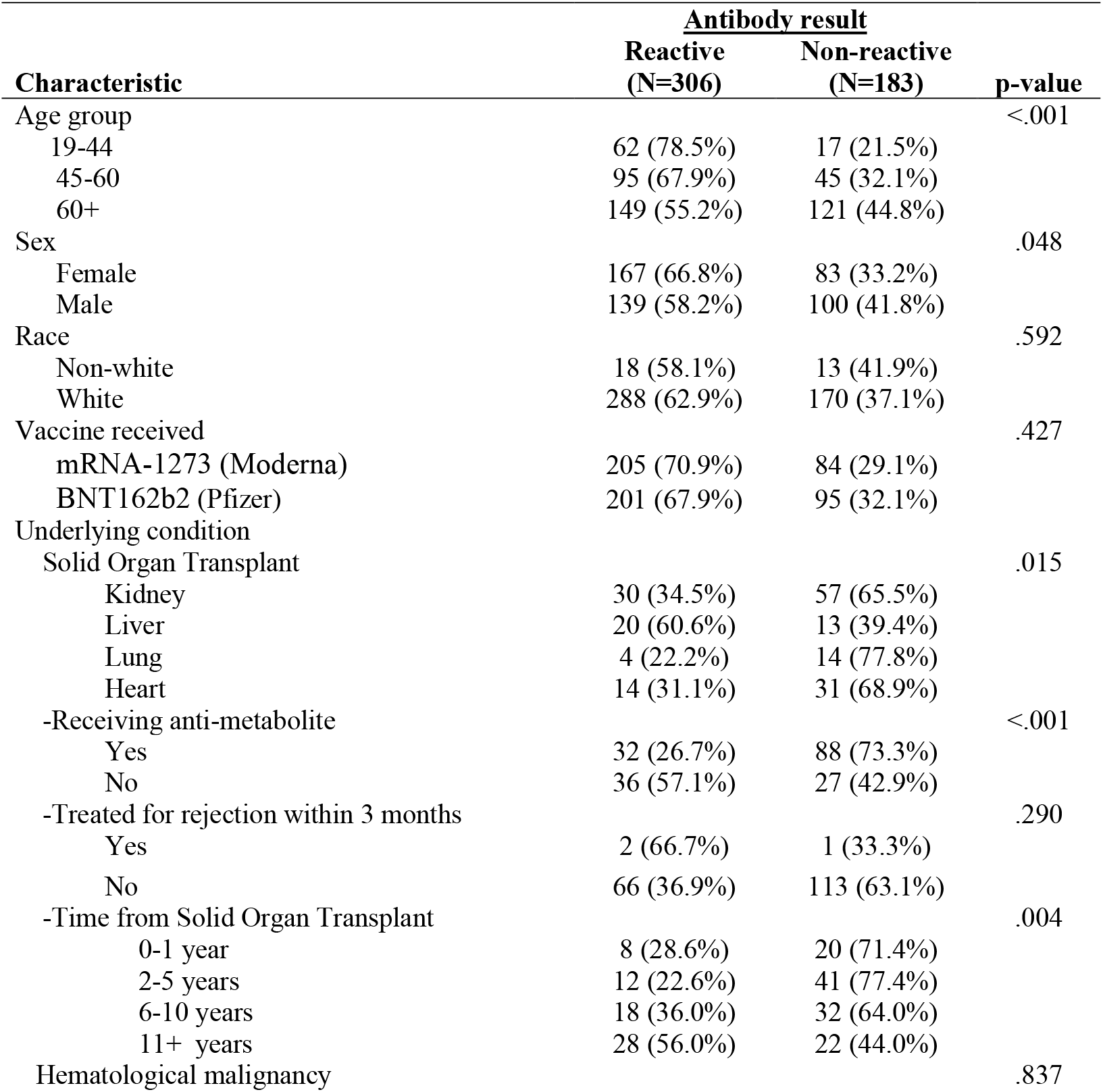
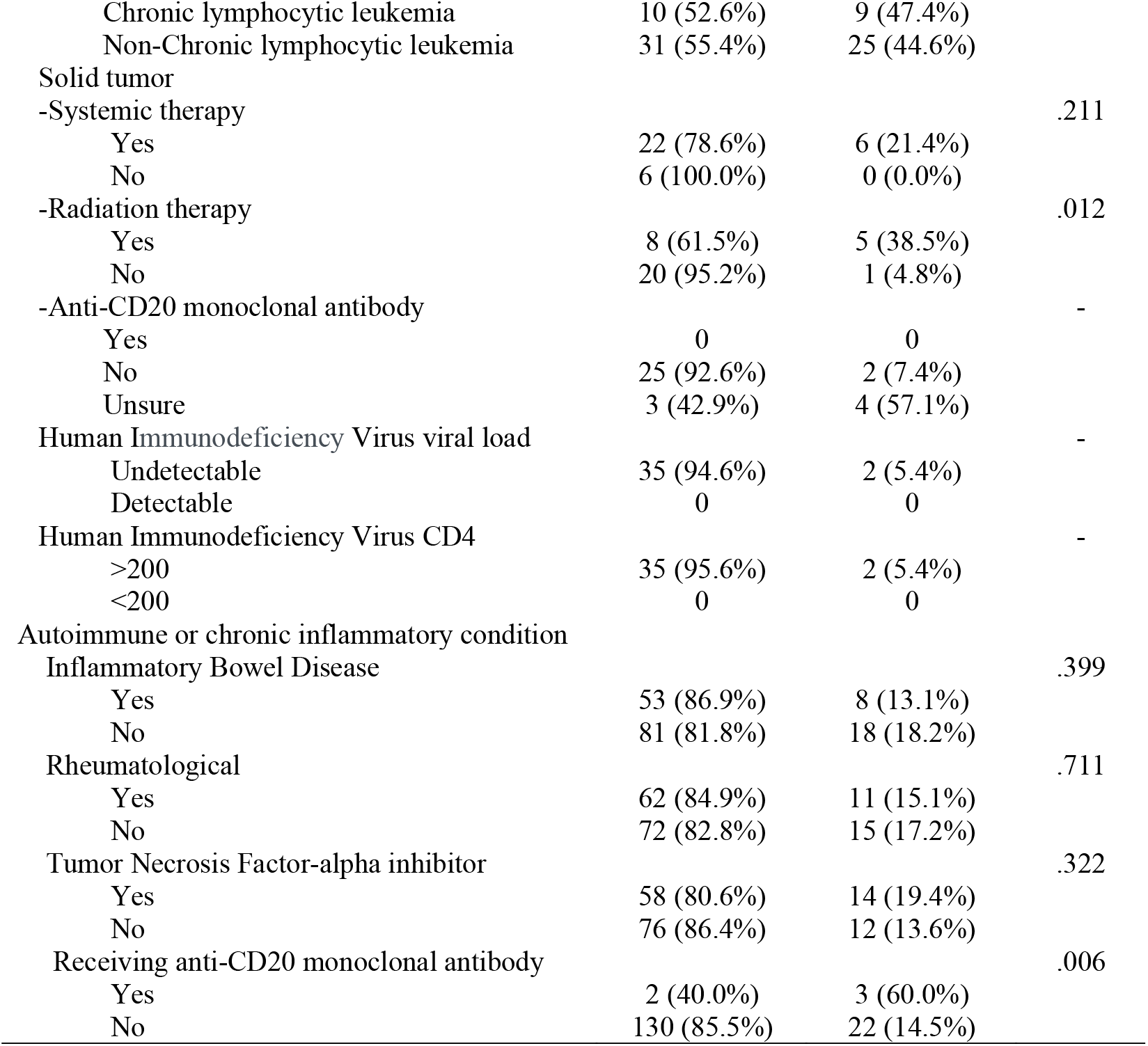
Clinical characteristics in immunocompromised patients by antibody testing results (N=489).

| Characteristic | Antibody result |  | p-value |
| --- | --- | --- | --- |
|  | Reactive<br>(N=306) | Non-reactive<br>(N=183) |  |
| Age group |  |  | <.001 |
| 19-44 | 62 (78.5%) | 17 (21.5%) |  |
| 45-60 | 95 (67.9%) | 45 (32.1%) |  |
| 60+ | 149 (55.2%) | 121 (44.8%) |  |
| Sex |  |  | .048 |
| Female | 167 (66.8%) | 83 (33.2%) |  |
| Male | 139 (58.2%) | 100 (41.8%) |  |
| Race |  |  | .592 |
| Non-white | 18 (58.1%) | 13 (41.9%) |  |
| White | 288 (62.9%) | 170 (37.1%) |  |
| Vaccine received |  |  | .427 |
| mRNA-1273 (Moderna) | 205 (70.9%) | 84 (29.1%) |  |
| BNT162b2 (Pfizer) | 201 (67.9%) | 95 (32.1%) |  |
| Underlying condition |  |  |  |
| Solid Organ Transplant |  |  | .015 |
| Kidney | 30 (34.5%) | 57 (65.5%) |  |
| Liver | 20 (60.6%) | 13 (39.4%) |  |
| Lung | 4 (22.2%) | 14 (77.8%) |  |
| Heart | 14 (31.1%) | 31 (68.9%) |  |
| -Receiving anti-metabolite |  |  | <.001 |
| Yes | 32 (26.7%) | 88 (73.3%) |  |
| No | 36 (57.1%) | 27 (42.9%) |  |
| -Treated for rejection within 3 months |  |  | .290 |
| Yes | 2 (66.7%) | 1 (33.3%) |  |
| No | 66 (36.9%) | 113 (63.1%) |  |
| -Time from Solid Organ Transplant |  |  | .004 |
| 0-1 year | 8 (28.6%) | 20 (71.4%) |  |
| 2-5 years | 12 (22.6%) | 41 (77.4%) |  |
| 6-10 years | 18 (36.0%) | 32 (64.0%) |  |
| 11+ years | 28 (56.0%) | 22 (44.0%) |  |
| Hematological malignancy |  |  | .837 |
| Chronic lymphocytic leukemia | 10 (52.6%) | 9 (47.4%) |  |
| Non-Chronic lymphocytic leukemia | 31 (55.4%) | 25 (44.6%) |  |
| Solid tumor |  |  |  |
| -Systemic therapy |  |  | .211 |
| Yes | 22 (78.6%) | 6 (21.4%) |  |
| No | 6 (100.0%) | 0 (0.0%) |  |
| -Radiation therapy |  |  | .012 |
| Yes | 8 (61.5%) | 5 (38.5%) |  |
| No | 20 (95.2%) | 1 (4.8%) |  |
| -Anti-CD20 monoclonal antibody |  |  | - |
| Yes | 0 | 0 |  |
| No | 25 (92.6%) | 2 (7.4%) |  |
| Unsure | 3 (42.9%) | 4 (57.1%) |  |
| Human Immunodeficiency Virus viral load |  |  | - |
| Undetectable | 35 (94.6%) | 2 (5.4%) |  |
| Detectable | 0 | 0 |  |
| Human Immunodeficiency Virus CD4 |  |  | - |
| >200 | 35 (95.6%) | 2 (5.4%) |  |
| <200 | 0 | 0 |  |
| Autoimmune or chronic inflammatory condition |  |  |  |
| Inflammatory Bowel Disease |  |  | .399 |
| Yes | 53 (86.9%) | 8 (13.1%) |  |
| No | 81 (81.8%) | 18 (18.2%) |  |
| Rheumatological |  |  | .711 |
| Yes | 62 (84.9%) | 11 (15.1%) |  |
| No | 72 (82.8%) | 15 (17.2%) |  |
| Tumor Necrosis Factor-alpha inhibitor |  |  | .322 |
| Yes | 58 (80.6%) | 14 (19.4%) |  |
| No | 76 (86.4%) | 12 (13.6%) |  |
| Receiving anti-CD20 monoclonal antibody |  |  | .006 |
| Yes | 2 (40.0%) | 3 (60.0%) |  |
| No | 130 (85.5%) | 22 (14.5%) |  |

Among hematological malignancy patients, there was no difference between seropositivity among chronic myelogenous leukemia (CLL) (52.6%, 10/19) versus non-CLL patients (55.4%, 31/56) (p=.837). In contrast, among patients with solid tumors, radiation therapy was associated with vaccine failure compared to radiation therapy (61.5%, 8/13 vs 95.2%, 20/21) seropositive, respectively (p=.012). Finally, use of an anti-CD20 monoclonal antibody within the prior 12 months was associated with failure to generate antibodies among patients with autoimmune or chronic inflammatory conditions, compared to no anti-CD20 monoclonal antibody (40.0%, 2/5 vs 85.3%, 128/150) seropositive, respectively (p =.007).

#### Antibody levels

Next, we analyzed IgG SAR-CoV-2 antibody levels across patient subgroups. While it appeared that levels were significantly lower in immunocompromised patients compared to HCW (**Figure 2A**), this finding was not surprisingly driven by the higher proportion of patients with negative antibody results in the immunocompromised group. When we excluded seronegative patients and analyzed only seropositive study participants **(Figure 2B)**, there were no statistically significant differences between antibody levels of seropositive healthy individuals (median [IQR] 7.2 [4.1,13.7]) and those of seropositive persons with autoimmune (median [IQR] 7.1 [3.0,13.4]), hematological malignancy (median [IQR] 10.5 [3.7,27.1]) and HIV (median [IQR] 8.2 [5.5,17.2]).

**Figure 2A.**
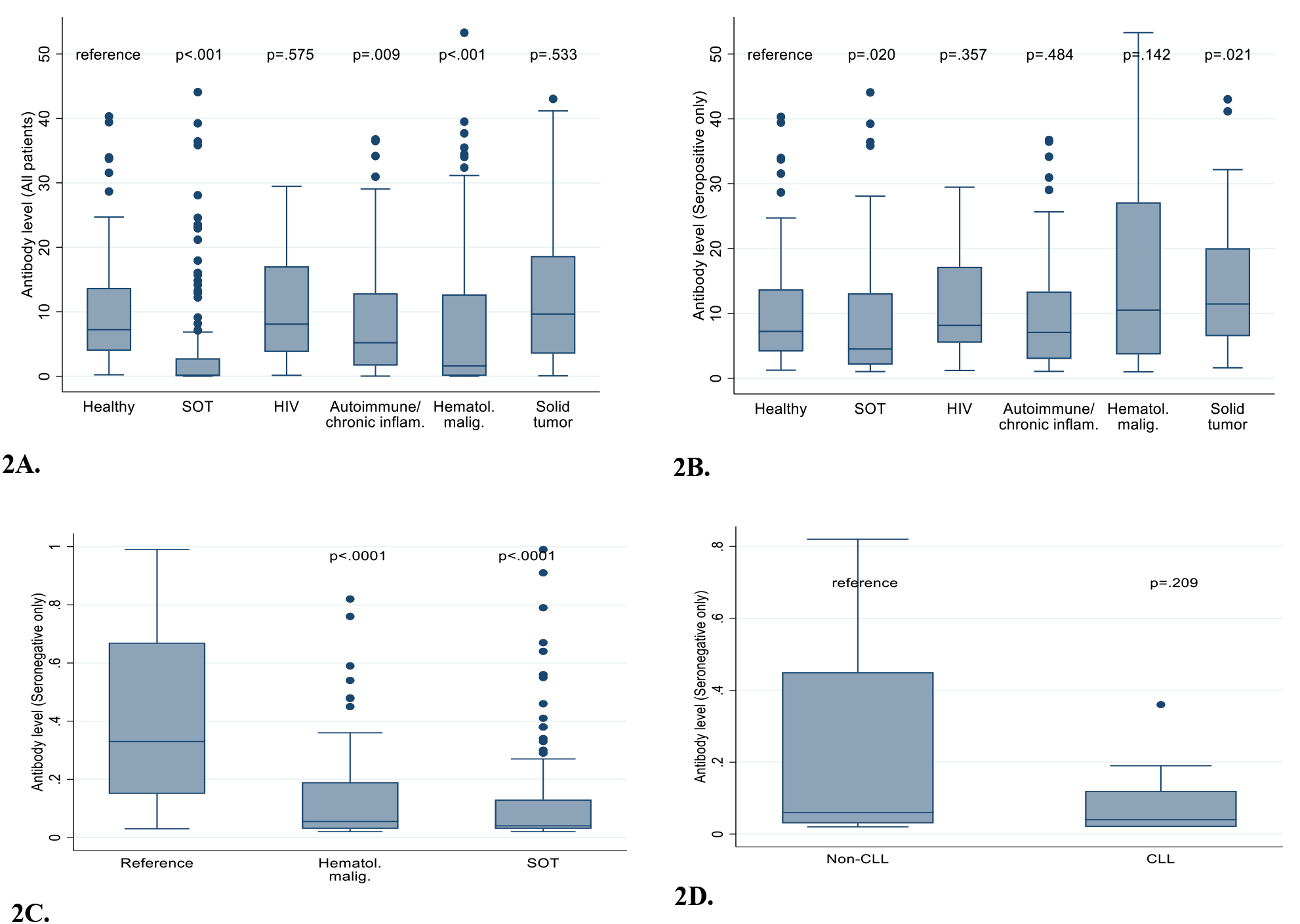
All antibody levels (seropositive and seronegative patients) in healthy healthcare workers and immunocompromised patients. **Figure 2B**. Comparisons of antibody levels among only patients with positive results. **Figure 2C**. Comparison of antibody levels among only patients with negative results, demonstrating that near absence of antibodies in many SOT recipients and patients with hematological malignancies. **Figure 2D**. Comparison of antibody levels CLL vs non-CLL hematological malignancy patients with negative results.

Solid tumor appeared to have a higher median antibody level when compared to the HCW (median [IQR] = 11.5 [6.5,20.1], p=.021). In contrast, antibody levels were significantly lower among SOT recipients compared to healthy controls (median [IQR] 0.2 [0.0,2.8], p=0.02). Since a recent report suggested that IgG responses vary even among individual with “negative” results,^20^ we compared antibody levels among patients with non-reactive or equivocal antibody levels (i.e., levels < 1) only (**Figures 2C** and **2D**). Overall, antibody levels from “seronegative” SOT (median [IQR] 0.0 [0.1,0.7]) and hematological malignancy (median [IQR] 0.1 [0.0,0.2]) patients were significantly lower than antibody levels from other seronegative patient groups (median [IQR] 0.3 [0.1,0.7], p <.001). Furthermore, antibody levels from seronegative CLL patients were not significantly lower than those of other patients with hematological malignancies (non-CLL median[IQR] = 0.1 [0.0,0.5]; CLL 0 [0,0.1], p =.2090). The effect of time from vaccination on antibody level was similar among HCW compared to immunocompromised patients, with a decline in −2.2 versus −1.1 units per 1 month from vaccination, respectively (p=0.158).

#### Pseudovirus neutralization assays

Neutralization titers were performed on a quasi-random subset of 66 participants (HCW=30, immunocompromised=36). Characteristics of these participants are shown in **Table 4**, and neuralization titers are shown in **Figures 3A** and **3B**. We observed a strong, positive correlation between antibody levels and neutralization titers (Spearman r = 0.91, p < 0.0001). Neutralization titers appeared to be lower among immunocompromised patients (median [IQR] = 33.0 [9.2,76.9]) compared to HCW (median [IQR] = 133.7 [69.6,340.6], p<.001), mirroring the lower antibody levels from this sample of immunocompromised patients (healthy median [IQR] = 8.2 [3.6,13.7] vs immunocompromised = 1.2 [0.6,9.1], p<.001).

**Table 4.**
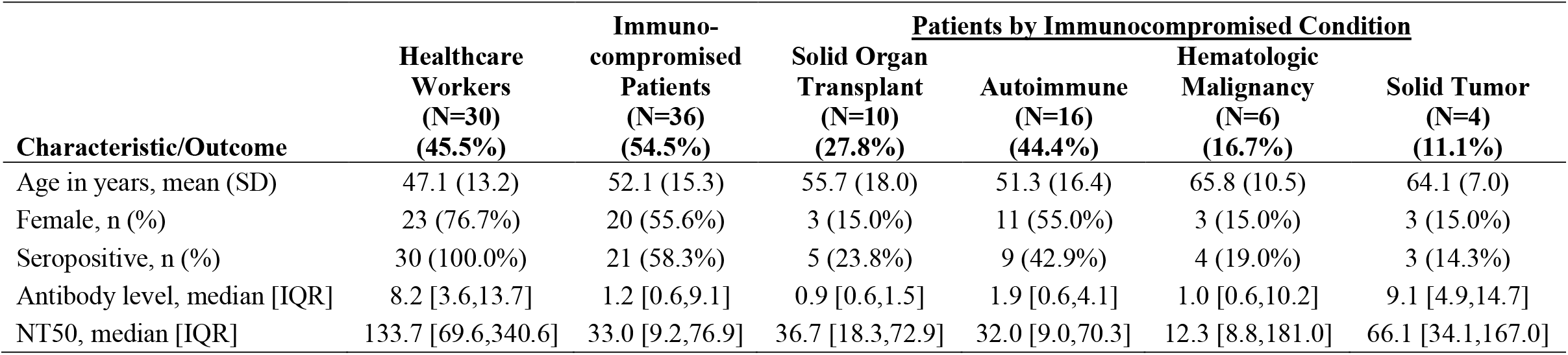
Characteristics and pseudovirus neutralization testing in healthcare workers and immunocompromised patients (N=66).

**Figure 3A.**
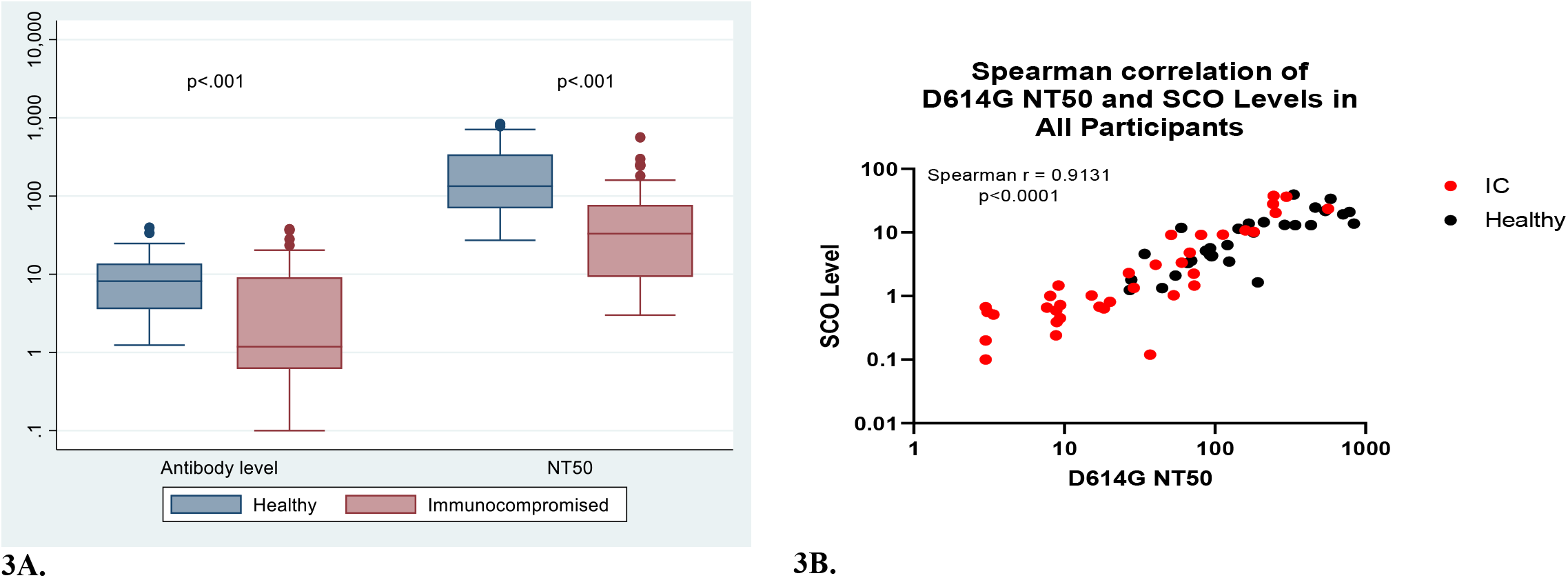
Comparisons of antibody levels and NT50 across the 30 HCW and 36 immunocompromised patients whose serum underwent pseudovirus neutralization testing. The NT50 was defined as the highest serum dilution that neutralizes >50% of the pseudovirus and antibody levels. **Figure 3B**. Scatter plot of D614G NT50 pseudovirus (x-axis) by Beckman Coulter extinction coefficients (y-axis). Black filled round dots depict health healthcare workers and red filled dots depict immunocompromised patients.

## DISCUSSION

In this prospective, longitudinal study of immunocompromised patients who have completed their COVID-19 vaccine series, we found that compared to healthy HCW, seropositivity was much lower in SOT recipients and patients with hematologic cancers (37.2% and 54.7%, respectively). In contrast, seropositivity among patients with solid neoplasms and autoimmune conditions approached those of HCW though remained lower (over 80% each versus 98.1%, respectively). Importantly, well-controlled patients with HIV mounted antibody responses nearly identical to those of healthy HCW. SARS-CoV-2 neutralization titers were generally proportional to antibody levels, though larger studies will be needed to fully assess whether subsets of immunocompromised patients fail to neutralize the virus. Finally, even among immunocompromised patients, some individuals mounted both antibody levels and neutralization capabilities that matched those of healthy HCW. Taken together, our findings demonstrate the heterogeneity of the humoral immune response to COVID-19 vaccines based on underlying immunosuppressive condition and highlight an urgent need to optimize and individualize COVID-19 prevention in these patients. Our findings also have implications on public health guidance, particularly given revised CDC recommendations permitting vaccinated individuals to abandon masking and social distancing in most settings.

We found several risk factors of vaccine failure in the different subgroups of immunocompromised patients, such as lung transplantation, use of antimetabolites or anti-CD20 monoclonal antibodies, and radiation therapy. Only 22.2% of lung transplant recipients in our cohort were seropositive, compared to 38.8% of other SOT recipients combined. Although this difference did not reach statistical significance, this observation remains biologically plausible, as lung transplant recipients generally receive higher doses of immunosuppressive medications than other organ transplant recipients and are considered to be at a much greater risk for infection. Similarly to a report by Boyarsky et al.,^21^ we found that patients who underwent their transplants within a year of vaccination were less likely to respond, a finding that is an indicator of a heightened state of immunosuppression in the early post-transplant period. Contrary to previous findings,^20^ among the participants with hematological malignancies, there was no statistically significant difference in the seropositivity of CLL versus non-CLL patients. Nonetheless, since circulating lymphocytes in patients with CLL are usually non-functional CLL cells, seronegative patients with CLL had much lower antibody levels than did other patients with hematological malignancies, suggesting true absence of an antibody response. Use of anti-CD20 monoclonal antibodies predicted vaccine failure in patients with autoimmune conditions, confirming that patients with B-cell aplasia cannot produce antibodies. Furthermore, radiation therapy was associated with failure to generate antibodies in patients with solid tumors, which is likely due to the toxic effects of radiation therapy on lymphocyte function.^22^ Finally, although it is extremely encouraging that 94% of participants with HIV responded to the vaccines, this group of patients continues to be a marginalized group of patients with poor access to vaccination,^23^ and outreach efforts should focus on increasing awareness of vaccination in these patients. Unlike a prior report,^21^ there was no association between type of mRNA vaccine and antibody response.

Despite the associations between specific immunosuppressive drugs and vaccine failure, we urge our patients not to self-discontinue these potentially life-saving medications. This is particularly true for organ transplant recipients, in whom the discontinuation of antimetabolites for the theoretical possibility of responding to vaccination will place the patient at risk for developing allograft rejection. This guidance is echoed by the American Society of Transplantation (AST), ^24^ which recommends against any modification in immunosuppressive medications at this time. Similarly, the National Comprehensive Cancer Network (NCCN) recommends the administration of COVID-19 vaccines when available, given the complete lack of data regarding whether vaccinating patients between chemotherapy cycles or during neutrophil recovery would have any beneficial impact on vaccine responses.^25^ In contrast, the American College of Rheumatology does recommend temporary interruptions in certain immunosuppressive medications (such as methotrexate, anti-CD20 monoclonal antibodies, and biologics) around the time of vaccination in patients with stable rheumatological diseases.^26^ Patients receiving such medications should discussion decisions to interrupt therapy with their providers.

Whether routine measurement of COVID-19 vaccine responses will become part of clinical practice remains to be seen. Multiple US societies such as CDC,^27^ Food and Drug Administration,^28^ AST,^24^ NCCN,^25^ and American Society of Hematology, Transplantation and Cellular Therapy^29^ currently recommend against routine assessment of antibody responses after vaccination outside of a study irrespective of the underlying immunocompromising condition. Additionally, immune correlates of protection, seroprotection titers, the association between neutralization and protection, and the contribution of T-cell responses to protection are not yet defined. For instance, although our seropositive immunocompromised patients (except SOT recipients) had antibody levels comparable to those of HCWs, neutralization titers appeared lower in some but not all immunocompromised patients. The clinical implications of these observations remain uncertain. Nonetheless, there is historical precedence for post-vaccine serological monitoring in immunocompromised patients, such as with hepatitis B vaccination, for which a booster series is recommended for non-responders.^30–31^ However, for many other vaccines such as influenza and herpes zoster, post-vaccine serological monitoring is not standard of care.^32,33^ Instead, immunocompromised patients are simply counseled that their risk of infection is higher than that of the general population.

There has been immense interest in re-vaccinating immunocompromised patients, but due to the regulatory landscape within the U.S. and the dearth of data on safety and efficacy, re-vaccination in the U.S. is not currently advised. In contrast, re-vaccination of immunocompromised patients, such as SOT recipients and patients receiving anti-CD20 monoclonal antibodies has been standard of care in France since the spring of 2021.^28^ Two recent studies from the U.S. and France of SOT recipients who did not respond to the initial vaccine series showed that 67% and 56% of patients also failed to respond to boosters, respectively.^34^ There was some evidence that response to the booster was associated with the initial post-vaccine antibody titer; indeed, our results suggest that some patients with non-reactive antibody levels may have some low-level antibody production compared to others (such as SOT recipients and CLL patients), in whom a negative result appears to indicate near absence of antibodies. Although we anxiously await the results of an ongoing randomized trial of revaccination in SOT recipients (NCT04885907),^35^ it is not surprising that many immunocompromised patients will never mount an antibody response. Should boosters and serological monitoring become routine care, patients who fail to respond to re-vaccination should be referred to clinical trials of other preventive measures such as monoclonal antibodies and prophylactic antivirals.

Limitations of this study include lack of longitudinal sampling and assessment of cellular immunity. Nonetheless, our data call for the need for funding to better understand immune responses to COVID-19 vaccines in immunosuppressed patients. Future studies should focus on correlating immune responses with clinical efficacy of COVID-19 vaccines and measurement of other correlates of immunity such as T-cell and memory B-cell responses, particularly in seronegative patients. There is a critical need to develop studies of revaccination, and for countries providing revaccination to publish their data. There is also a critical need to design trials of passive immunity using monoclonal antibodies or direct-acting antivirals for the prevention of COVID-19 in immunocompromised patients. Finally, results such as ours should not be used to fuel vaccine hesitancy, but rather to encourage vaccination and emphasize the need for ongoing vigilance until additional interventions are available.

## Data Availability

Data is not available from this ongoing study.

## Acknowledgements

The authors thank the following individuals for providing multidisciplinary commitment to advance scientific discovery and quality of patient care in response to the COVID-19 pandemic: Jordan Bartlow, Logan Baylor, Sarah Behr, Shannon Buono, Vibha Chauhan, Maddie Chrisman, Shawna Chylinski, Cathy Cochran, Lindsay Coughenour, Nicole Czolba, Celeste Duprey, Kelly Friday, Megan Fritz, Kailey Hughes, Trevor Katich, Kristin Kerfoot, Joshua Kohl, Elaine Lander, Michelle Lucas, Aimee Majeski, Traci McGaha, Rachel McGargle, Audrey Paul, Amber Shaffer, Jordan Shayer, Lisa Sheehan, Kristin Shoemaker, Lori Snyder, Courtney Starrett, Abbey Sung, Christina Tedesco, and Jamie Voyten.

---

**Supplemental Table 1.**
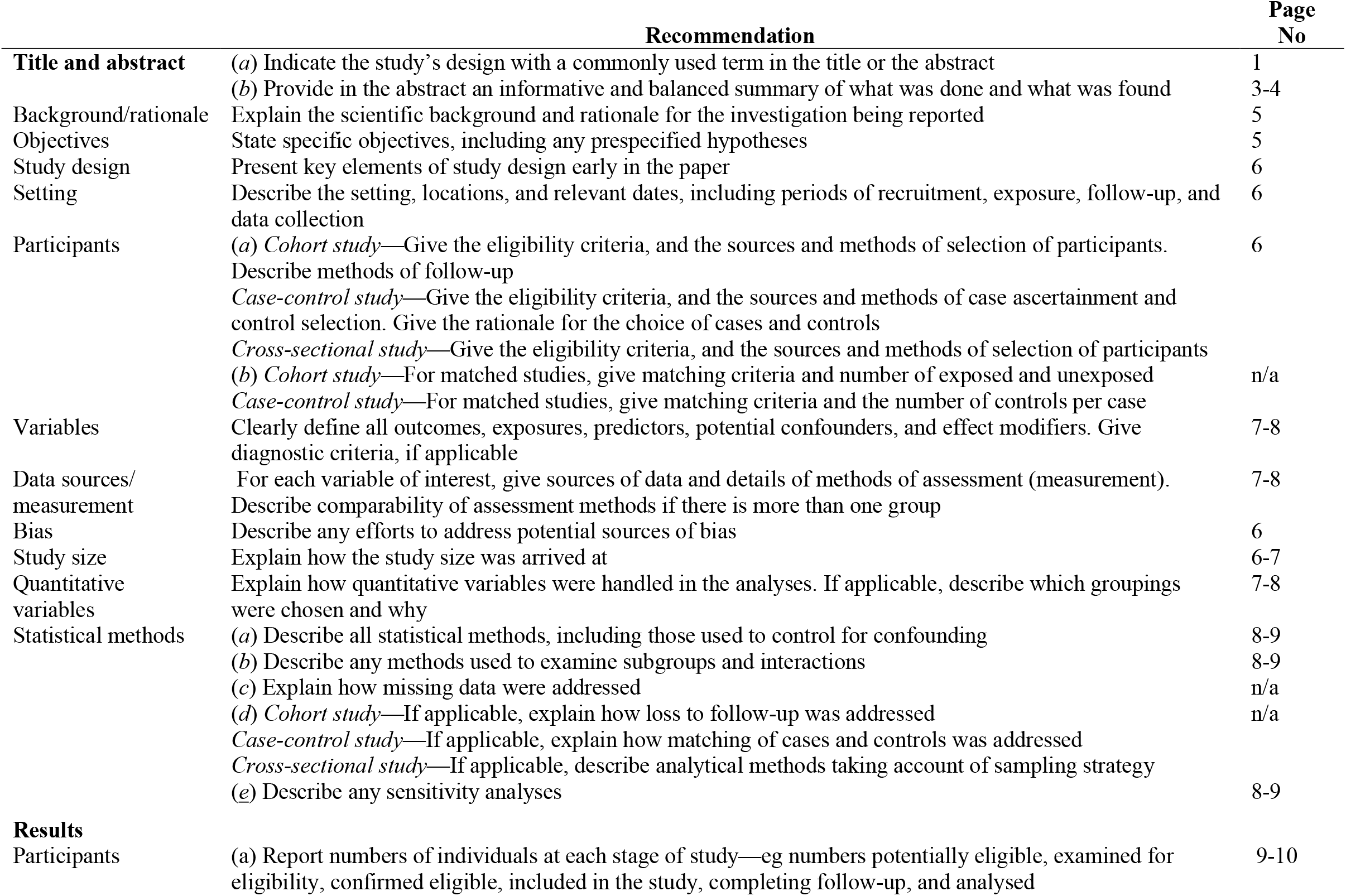
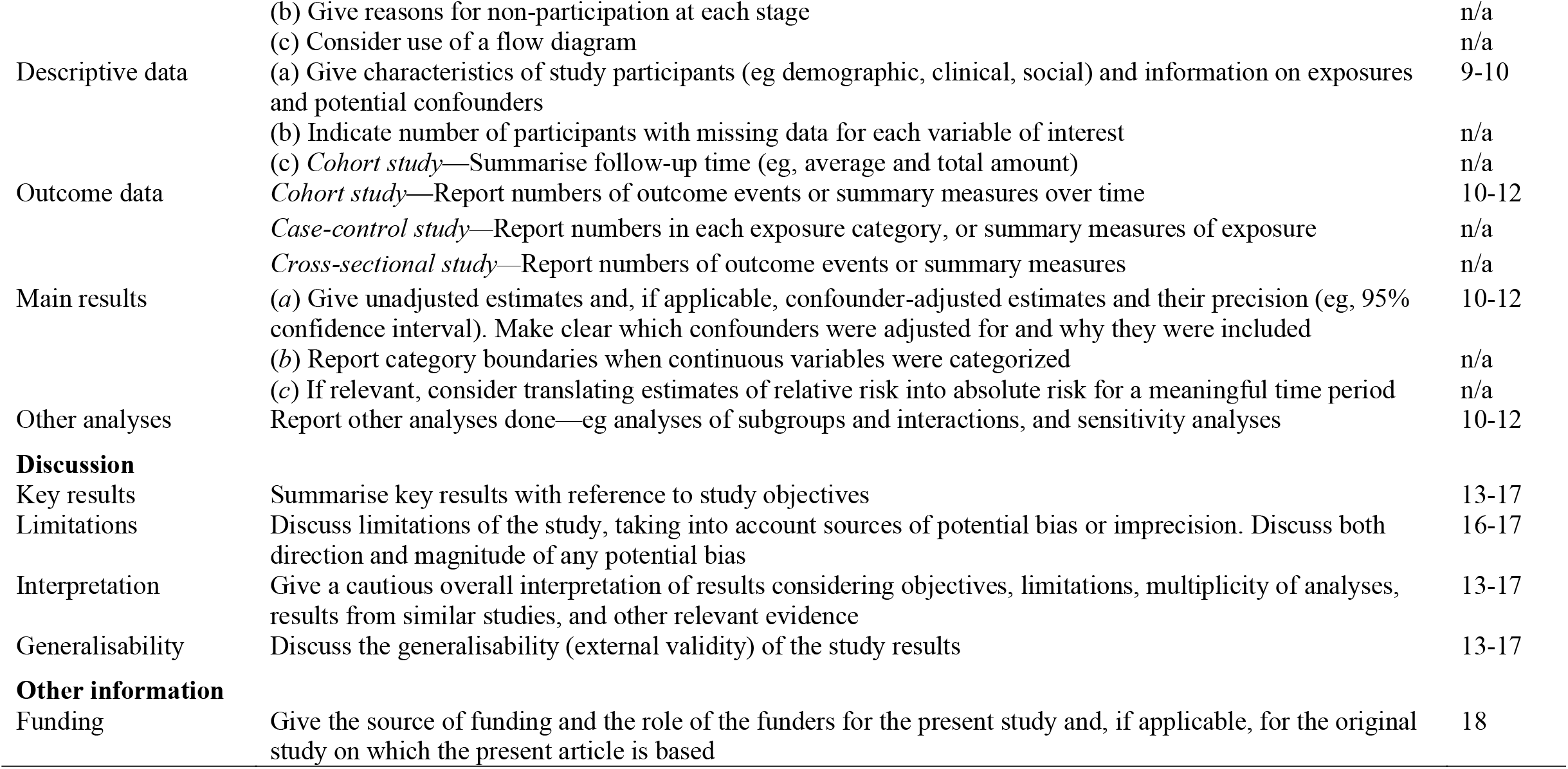
The Strengthening the Reporting of Observational Studies in Epidemiology (STROBE) Checklist.

